# Impact of a novel comprehensive empathy curriculum at Leicester Medical School: Protocol for a longitudinal study

**DOI:** 10.1101/2024.02.08.24302205

**Authors:** Leila Keshtkar, Amber Bennett-Weston, Andy Ward, Rachel Winter, Simon Gay, Jeremy Howick

## Abstract

**Background:** Empathy appears to decline throughout medical school and is highly variable among qualified healthcare practitioners. To address these problems, the Leicester Medical School has designed an enhanced empathy curriculum that runs throughout all five years of medical school. The impact of this curriculum must be measured.

**Objective:** To evaluate the impact of the enhanced empathy curriculum using qualitative and quantitative data.

**Study design:** Prospective longitudinal study

**Setting:** Leicester medical school

**Participants:** All students (from year one to five) at the Leicester Medical School will be eligible for participation. There are currently approximately 300 students in each of the five years of medical school and we anticipate that we will recruit approximately 100 medical students per medical school year for the quantitative data (total of 500 students) and approximately 10 students per medical school year for the qualitative data (total of 50 students).

**Data collection:** We will annually collect the following data for a period of seven years to ensure a comprehensive dataset for three full cohorts of medical students. The main reason for selecting the seven-year timeframe is that the empathy curriculum recently started in medical school (2022-2023 academic year), and it takes a few years for it to stabilise and yield meaningful results.

1. Student empathy levels (for example, using the student version of the self-reported Jefferson Scale for Empathy (JSE-S) and the observer-reported Consultation and Relational Empathy (CARE) measure).
2. Satisfaction with the empathy curriculum (using routinely collected student survey data).
3. Satisfaction with the overall medical school curriculum (using routinely collected student survey data).
4. Student experiences of the empathy curriculum (using qualitative student interviews with a purposive sample of five to 10 medical students from each year).
5. Student well-being data (for example, the frequency and severity of well-being issues).

**Reporting elements:** We will report on:

1. Number of individuals at each stage of the study.
2. Descriptive data that includes (student characteristics and potential confounders).
3. Outcome data (empathy levels, student satisfaction with curriculum, student experience). We will also report on the relationships between these data (for example between empathy levels and student satisfaction with curriculum).

**Dissemination:** Findings will be disseminated through international conferences, news and peer-reviewed journals.

**Funding:** This study is funded by the Stoneygate Trust. The funder had no role in any part of the development or writing of this paper.

**Ethics:** The project and associated documents were approved by the University of Leicester Research Ethics Board (Ethical approval number: 42404-lk252-ls:medicine).

## 1. INTRODUCTION

### 1.1. Background and rationale

Enhanced empathic care has several benefits for patients ranging from reducing their pain to improving their quality of life.^1^ Empathic care also appears to benefit practitioners by improving their well-being and reducing burnout,^2,3^ while improving their sense of professionalism and job satisfaction.^4^ This is especially important since medical student mental health leaves much to be desired. A systematic review of 16 studies (5728 students) found that an average of 27.2% of medical students had depressive symptoms, and on average 11.1% of medical students experienced suicidal ideation.^5^ The same study found that these rates were higher than they were before the same students entered medical school. Relatedly, a large study with over 45,000 participants found that rates of burnout were higher among medical students and residents than in the general population.^6^ Randomised trials show that training improves healthcare practitioner empathy.^7^

Unfortunately, the extent to which patients report their practitioners to be empathic varies widely,^8^ and medical student empathy appears to decline throughout medical school.^9,10,11^ To encourage the benefits of empathic care for patients and practitioners, Leicester Medical School developed and implemented a novel empathy curriculum throughout all five years of medical school. The curriculum includes five streams:

1. “Walk a mile in your shoes” and creative health. In this curriculum stream, students are provided with vicarious experiences (such as wearing an ageing suit) and then are mentored to express their experience creatively (for example through a poem, story, painting, or play).
2. Getting real patients into the lecture theatre during the teaching of pathophysiology.
3. Empathy-focused communication skills training.
4. Wellbeing.
5. A “transition course” helping students transition from the pre-clinical to clinical phases of medical school.

Each of these curriculum streams is being evaluated individually. It is also important to evaluate the overall impact of the curriculum.

## 2. RESEARCH QUESTIONS AND OBJECTIVES

The overarching aim of this longitudinal study is to evaluate the impact of the enhanced empathy curriculum and answer the following questions:

1. Does the empathy curriculum positively impact the decline in medical student empathy?
2. What are the levels of student satisfaction with the empathy curriculum?
3. What are student experiences of the empathy curriculum?

## 3. METHODS

### 3.1. Study design

A longitudinal study that measures the impact of the empathy curriculum using both quantitative data (about empathy levels, student satisfaction with the empathy curriculum and satisfaction with the overall medical school curriculum) and qualitative data (about student experiences with the medical school curriculum).

### 3.2. Setting

Leicester Medical School.

### 3.3. Data collection

We will annually collect data for a period of seven years on empathy levels, student satisfaction with the curriculum, student experience of the curriculum, and student well-being.

#### 3.3.1. Empathy levels

We will administer empathy questionnaires. These will include, for example, the self-reported JSE-S,^12^ and where feasible and appropriate, patient-reported empathy data using, for example, the CARE measure.^13^

#### 3.3.2. Student satisfaction with empathy curriculum

Student satisfaction data will include satisfaction with the empathy curriculum and satisfaction with the overall medical school curriculum. These data (i.e., satisfaction with the empathy curriculum, and satisfaction with the overall medical school curriculum) will be collected through questions that are embedded in the medical school feedback survey to students in all five years of medical school.

#### 3.3.3. Student experience with empathy curriculum

We will explore the students’ views and experiences of the medical school empathy curriculum by conducting qualitative interviews. We will purposively select a group of five to 10 students per medical school year who have high, low, and mid-range JSE-S scores to explore their views and experiences of the medical school empathy curriculum (A list of questions provided in Supplement File 1). Student selection will be based on their expressed interest in participating in the interviews during the empathy level data collection stage.

#### 3.3.4. Student well-being data

We will collect data regarding the prevalence and severity of well-being incidents from the pastoral support unit.

### 3.4. Participants and eligibility

#### 3.4.1. Empathy levels

All students at the Leicester Medical School will be eligible for participation. For the JSE-S, first and second-year medical students will be approached during one of the compassionate holistic diagnostic detectives (CHDD) teaching sessions. Third-fourth- and fifth-year medical students will be approached during their study days in the medical school.

For CARE, phase II students may be asked to give the CARE measure to patients they treat on their placements.

If other validated measurement tools are developed, we will consider using them. To ensure continuity of data, we will either use these in addition to the JSE-S and CARE or ensure that they are sufficiently correlated with those scales.

#### 3.4.2. Student satisfaction with empathy curriculum

All students at the Leicester Medical School are routinely invited to fill in a student survey at the end of each medical school year (This survey is administered by the Leicester Medical School).

#### 3.4.3. Student experience of the medical school curriculum

All students at Leicester Medical School will be eligible for participation. Participants will be selected using a purposeful, maximum variation sampling strategy.^14,15^ Purposive sampling involves the researcher using their knowledge of the literature, the research setting, and the research questions to strategically select participants from whom we can learn a great deal about the phenomenon under investigation.^14,15^ Specifically, a maximum variation approach involves selecting participants who can offer a range of perspectives on dimensions of relevance to the research questions.^16^ Accordingly, we will purposefully select medical students from each year group who have high, low, and mid-range JSE scores. The selection of these students will be based on their expressed interest in participating in the interviews during the empathy level data collection stage to explore their views and experiences of the medical school empathy curriculum. Only students who provide their consent to participate will be included in the interviews at the end of each year of medical school.

### 3.5. Sample size

#### 3.5.1. Empathy levels

There are approximately 300 new medical students every year at the Leicester Medical School. We will have an active recruiting strategy that involves encouragement from the head of the medical school and all teaching leads. On this basis, we anticipate that we will recruit 100 medical students in each year of medical education.

For the CARE measure, we will ask the purposefully selected medical students with high, low, and mid-range JSE scores to get their patients to fill in CARE questionnaires. Twenty-five patients per practitioner is believed to offer a reliable estimate, however, this is not feasible for medical students who do not treat as many patients as practitioners. We will therefore aim for 25 patients per group (low, medium, and high JSE scores).^17^ We will explore the feasibility of comparing CARE and JSE scores, to investigate correlations between the JSE and CARE measures.^18,19^ Achieving this will depend on ethical approval (students will have to agree for their JSE scores to be made available).

#### 3.5.2. Student satisfaction with the empathy curriculum

There are more than 300 students in each year of medical school (year one to five) and we anticipate that we will recruit more than 100 medical students in each year of medical education (a total of more than 500 students in each year of study).

#### 3.5.3. Student experience of empathy curriculum

We will aim to recruit a group of five to 10 medical students in each year of medical education (a total of 50 students in each year of study). We will cease recruitment and data collection when saturation is reached. Therefore, the exact sample size will be determined by the point at which we fully understand students’ views and experiences of the empathy curriculum at Leicester Medical School and no new ideas are being generated through the interviews.^20^

### 3.6. Ethics

The project and associated documents were approved by the University of Leicester Research Ethics Board (Ethical approval number: 42404-lk252-ls:medicine).

#### 3.6.1. Empathy levels

For JSE-S data collection, students will be provided with an information sheet (See Supplement File 2) and an informed consent form (See Supplement File 3) to sign, upon being invited to take part in the study. Only students who provide their consent to participate will be included in the study, and students will have the ability to remove their consent at any time during the study. They will also receive a privacy notice sheet (See Supplement File 4), which will provide information about how the University of Leicester collects and uses personal information when participants take part in this research project.

For the CARE measure, patients will receive an information sheet (See Supplement File 5) and a consent form (See Supplement File 6) to sign when invited to join the study. Participation is only for those who provide their consent to participate will be included in the study, and they can withdraw at any time. They will also receive a privacy notice sheet (See Supplement File 4).

#### 3.6.2. Student satisfaction with empathy curriculum

This survey is administered by the Leicester Medical School and no additional permission is required for this.

#### 3.6.3. Student experience with empathy curriculum

Those students who agree to participate will be given a participant information sheet detailing the consent process and more information about the interview (See Supplement File 7). They will also receive a privacy notice sheet (See Supplement File 4). They will subsequently be asked to sign a consent form (See Supplement File 8) before contributing to the study. Only students who provide their consent to participate will be included in the study. Participation will be voluntary, and students will have the ability to remove their consent at any time during the study. The anonymity of the participants and the confidentiality of data will be always maintained. The data will also be analysed anonymously such that the results are non-traceable to an individual participant.

## 4. MEASURES

### 4.1. Empathy levels

#### 4.1.1. Self-reported empathy, the Jefferson Scale

The Jefferson Scale^21^ is a self-rated scale with 20 items measured on a seven-point Likert scale. Possible scores range from 20 to 140, and higher scores are associated with greater degrees of empathy.

#### 4.1.2. Patient-reported empathy scale, the CARE measure

The CARE measure involves 10 questions answered on a five-point Likert scale and possible scores range from 10 to 50.^22^ Higher scores indicate higher levels of empathy.

#### 4.1.3. Other measures of empathy

If other evidence-based and validated measures will be developed we will consider them.

### 4.2. Student satisfaction with empathy curriculum

#### 4.2.1. Satisfaction with empathy curriculum

Satisfaction with the empathy curriculum will be assessed through student ratings of core components of the empathy curriculum by answering these questions:

1. To what extent are you satisfied with the empathy curriculum at Leicester Medical School? Response options include: very satisfied, somewhat satisfied, neither satisfied nor dissatisfied, somewhat dissatisfied, very dissatisfied.
2. Do you agree with this statement “The empathy curriculum this year will help me to become an empathic doctor”? Response options include: strongly disagree, disagree, neither agree nor disagree, agree, strongly agree.

#### 4.2.2. Satisfaction with the overall medical school curriculum

Student satisfaction with the overall curriculum will be taken from student ratings of the medical school curriculum.

### 4.3. Student experience of empathy curriculum

Qualitative data will be collected using semi-structured interviews. Students will receive an information sheet detailing the consent process and more information about the interview (See Supplement File 7). As is advocated under a qualitative approach, the questions included in the topic guide are broad and open-ended to allow space for participants to construct the meaning of the research phenomenon and to share as much, in their own words, as possible.^23^ The questions are designed to explore students’ understanding of the empathy curriculum, what they liked and disliked about the teaching they received as part of this curriculum and their suggestions for improvement. The topic guide will be piloted on a small sample of medical students (n=3) to ensure the questions are clear and comprehensive.^24^ After each interview, the topic guide will be reviewed. Supplementary questions will be added, if necessary, to allow exploration of interesting ideas in subsequent interviews.^23^

### 4.4. Student well-being

Student well-being will be assessed by collecting data on the frequency and severity of well-being incidents from the pastoral support unit.

## 5. PROCEDURE

### 5.1. Empathy levels

The online JSE-S will be administered to students during year-specific protected teaching time. Participant information and consent forms will be integrated into the online surveys. Registers will not be taken in the sessions when the surveys will be administered, so completion rates are based on the numbers in the whole year group, rather than on those present at the time of the surveys. Except for the voluntary option to enter email for receiving feedback, no personal identification information will be solicited. All individual data will be anonymised.

To compare the impact of the curriculum, we will conduct baseline measurements with all questionnaires with medical students (all years) at the outset of 2023.

For CARE, phase II students will be asked to give the CARE measure to patients they treat on their placements.

For other empathy measures that might be taken, we will integrate the procedure into the existing ones listed above.

### 5.2. Student satisfaction with the empathy curriculum

This data will be collected each year of the project by a feedback survey in medical school. All data will be completely anonymised to conserve confidentiality. This will be achieved by coding the data.

### 5.3. Student experience with empathy curriculum

This study will use a qualitative semi-structured interview, where the data source is individual interviews.

#### Sample size

The sample size differs depending on what the focus of the research is, what will have credibility and what resources are available.^25^ According to Creswell (2007),^26^ semi-structured interviews require a minimum sample size of between five and twenty-five. Saldana (2011),^27^ also identified that based on selected methodologies, a minimum of ten to thirty participants is needed to ensure more credible and trustworthy findings. Furthermore, the sample size is determined when saturation is reached and no new perspectives and insights are being found.^20^ Therefore, for this study, we will recruit a minimum sample of 50 participants to ensure richness in data to cover all or the majority of possible perspectives. The exact sample size will be determined by the point at which saturation is reached. The interviewees will be purposefully selected using a maximum variation sampling strategy from among medical students each year. Interview questions are designed to be broad and open-ended to explore students’ views and experiences regarding the empathy curriculum at Leicester Medical School.

#### Pilot testing

It is strongly recommended that interview topic guides be tested before being administered. We will achieve this by running a pilot study using a copy of the topic guide on a small sample of respondents that has the same characteristics as the intended sampling frame.^24^

#### Interview administration

To conduct the interviews, all the participants will be approached by email and receive a participant information sheet describing the content and the aim of the study (See Supplement File 2). This is intended to familiarise respondents with the project and provide them with background information about the topics that will be discussed during the interview. The respondents will have the opportunity to ask any questions to gain more information and clarify their understanding of the interview. Informed consent will be obtained before commencing interviews. The interviews will be pre-scheduled and will take place either face-to-face or online to accommodate different year groups of medical students (for example fifth-year medical students who might have left Leicester at the time of the study). All interviews will be semi-structured to discuss participants’ views and experiences of the empathy curriculum. The topic guide will be used to steer the discussion between interviewer and interviewee and ensure it remains on topic. However, the interviewer will be responsive to the participant’s developing accounts and therefore may pursue additional lines of inquiry.^28^ To avoid response bias, the interviewer will avoid using leading questions and will not express a personal opinion on any of the matters the participant might discuss.^29^ A list of interview questions is provided in Supplement File 1.

## 6. ANALYSIS

### 6.1. Empathy levels and student experience with empathy curriculum

We will use Analysis of variance (ANOVA) and an independent t-test to examine changes in the JSE-S and CARE scores. Pearson’s correlation will be used for the analysis of the association between patients’ overall satisfaction and perceived empathy of physicians.

We will analyse trends in all main variables and the interaction between variables through linear or non-linear interaction effects. Here is a list of main variables:

- JSE-S scores (continuous): represent the empathy levels of students measured by JSE-S.
- CARE scores (continuous): represent another measure of healthcare practitioners’ empathy provided by patients.
- (If obtained) other measures of student empathy.
- Overall satisfaction of empathy curriculum (categorical scale): measures students’ satisfaction with the empathy curriculum using a categorical scale, likely assessing levels of satisfaction (e.g., very satisfied, satisfied, neutral, dissatisfied, very dissatisfied).
- Satisfaction with the overall medical school curriculum (categorical scale).

And here are the hypothesized relationships that will be explored, subject to data availability:

- Relation between JSE-S and CARE measure (i.e., empathy level measures).
- The overall satisfaction of the empathy curriculum.
- Relation between empathy level measures and academic performance.
- Relation between the empathy level measures and students’ well-being.
- Relation between satisfaction with the empathy curriculum and the relation with students’ well-being.

Additional relationships will also be considered on an ad hoc basis.

### 6.2. Student satisfaction with the empathy curriculum

Qualitative data will be transcribed verbatim and uploaded into NVivo software to be analysed using reflexive thematic analysis. Reflexive thematic analysis is a six-phase method for “identifying, analysing and interpreting patterns of meaning (‘themes’) within qualitative data.”^30^

#### Phase 1: Familiarisation

The transcripts of the individual interviews will be read and re-read to facilitate the researcher’s familiarity with the dataset.

#### Phase 2: Coding

A series of codes will be developed and assigned to extracts from the transcribed text. In this process, codes are not pre-assigned. Concepts, interesting points and conversations among responses to the interview questions will be identified and assigned a code (a label that captures what is analytically interesting about the data.

#### Phase 3: Generating initial themes

Next, the codes will be compared with each other and similar codes will be highlighted and classified. Codes with common content then will be renamed, combined and a unified code will be provided. These unified codes will become candidate themes.

#### Phase 4: Developing and reviewing themes

In this phase, the emphasis will be placed on the development of more specific categories with related and similar contents. The candidate themes will be checked against the collated coded data extracts from which they were developed to ensure they represent a clear and coherent pattern of meaning. The entire qualitative dataset will be revisited to provide opportunities to re-code data in light of the developing analysis. Information that does not align with the study’s research questions will be removed, and main sub-themes and categories will be compared and contrasted to identify overarching themes. Thematic maps will be developed to explore the relationships between overarching themes and sub-themes, highlighting areas requiring further development (for example, themes that overlap).

#### Phase 5: Refining, defining and naming themes

After several iterations of the previous phase, key themes and subthemes from the data will be refined and named. Short summaries will be produced to describe each theme and these will be organised into a coherent analytic narrative.

#### Phase 6: Writing up

The themes and subthemes will be reported, accompanied by extracts of data that support the analytic narrative.

### 6.3. Predictors

We anticipate that changes in empathy (self-reported and patient-reported) and satisfaction with the empathy curriculum predict well-being. The basic model we hypothesize is the following:

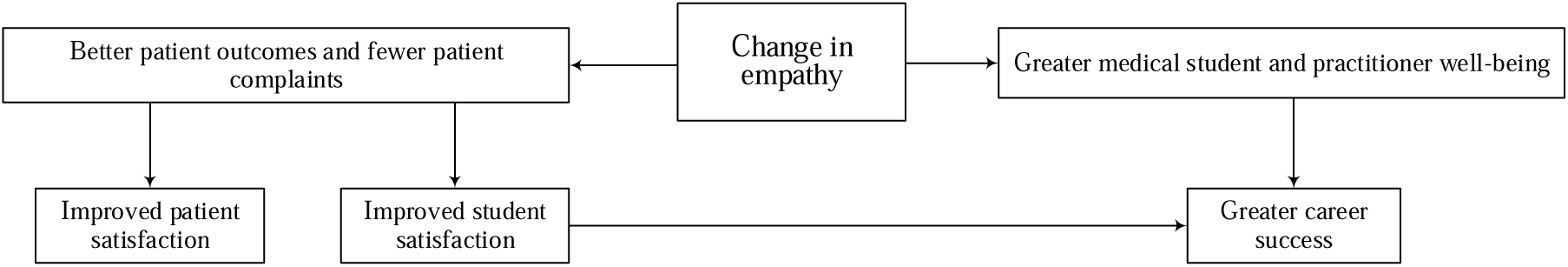

### 6.4. Potential confounders

There are potential confounders in this study that are inherent in the collection of observational data. For example, ‘Hawthorne Effects’ (i.e., participants’ behaviour changes due to their awareness of being part of a long-term study), different changes to the medical school curriculum and societal changes (e.g., pandemic diseases) that may affect the results.

To address Hawthorne Effects, we will administer questionnaires such as JSE-S to gather data without direct interaction. We will also use anonymised data and mention it during our data collection procedures through each year to reduce their awareness of being observed.

Changes introduced to the Leicester Medical School Curriculum could also modify the effect of the empathy curriculum over time. To address this, any modifications will be documented, and we will provide a detailed report and clarification about these changes. We will also conduct an annual audit of the medical school curriculum to identify potential modifiers and assess their potential relevance.

To address societal changes that may affect the curriculum (e.g., pandemic diseases), online sessions, online platforms and related strategies established by medical school will be used.

### 6.5. Bias

Bias is a fundamental concept that can affect many aspects of research. It refers to any trend or deviation from the truth in data collection, data analysis, interpretation and publication which can cause false conclusions.^31^ Understanding bias, its sources and its impact is crucial for informed and fair decision-making and rigorous research.

#### 6.5.1. Potential bias in measuring empathy levels

All the self-reported assessments are subject to several reporting biases including self-report bias, social desirability bias, recall bias, misclassification, measurement error bias, and confirmation bias. We will adjust for these using a variety of methods depending on the outcome measured.^32^

#### 6.5.2. Potential bias in measuring student satisfaction with the empathy curriculum

The medical school feedback survey as a self-reported assessment is also subject to several reporting biases. These biases including self-report bias, social desirability bias, recall bias, misclassification, measurement error bias, and confirmation bias should be considered.

#### 6.5.3. Student experience with empathy curriculum

We will consider different types of bias including interviewer-participant relationship, social desirability bias, response bias and sampling bias that may affect the data collection through interviews.^32,33^ Interview data are socially constructed through interviewer-participant interaction.^34^ Moreover, as part of the qualitative data analysis, the researcher(s) will be interpreting and making sense of participants’ views and experiences.^23^ These interpretations will be shaped by the researchers’ backgrounds, views and experiences.^35^ Therefore, it is essential that the researcher(s) remain mindful of their influence over data collection and analysis, by engaging in reflexivity.^23^ Reflexivity is an “immediate, continuing, and dynamic self-awareness” requiring the researcher to critically reflect upon their position within and impact upon, the research process.^36^

## 7. RESULTS AND REPORTING ELEMENTS

We will report on the elements listed below.

1. Numbers of individuals at each stage of study (e.g., numbers potentially eligible, examined for eligibility, confirmed eligible, included in the study and analysed).
2. Descriptive data that includes:

- Characteristics of study participants (e.g., clinical, social) and potential confounders (e.g., age, gender, cultural background)
- Number of participants with missing data for each variable of interest
3. Outcome data:

Empathy levels

- Average JSE-S scores and where feasible CARE scores for students in specific years and comparing it across different years of medical education
- Relation between JSE-S and CARE measure
- If further empathy measures are developed, these may also be included in the analysis; if so, they will not replace the other measures, and correlations between the new and established measures will also be reported.
Student satisfaction with empathy curriculum

- The overall satisfaction of the empathy curriculum by answering these two questions, which will be embedded in the medical school feedback survey:

1. Do you agree with this statement “The empathy curriculum this year will help me to become an empathic doctor”? Response options include: strongly disagree, disagree, neither agree nor disagree, agree, strongly agree.
2. Do you agree with this statement “The empathy curriculum this year will help me to become an empathic doctor”? Response options include: strongly disagree, disagree, neither agree nor disagree, agree, strongly agree.
Student experience with empathy curriculum

- Exploring the students’ views and experiences of the empathy curriculum and main themes and sub-themes will be generated through reflexive thematic analysis of interview data, accompanied by extracts from individual interviews.
4. Statistical analyses

- Changes in the JSE-S related to the year of medical school, gender and speciality interest.
- Trends and relationships between the empathy level, student satisfaction with the empathy curriculum and student experience with the empathy curriculum.
- Unadjusted estimates and, if applicable, confounder-adjusted estimates and their precision (e.g., 95% confidence interval). The confounders include: different changes to the medical school curriculum and societal changes (e.g., pandemic diseases).
- Category boundaries when continuous variables.
- Other analyses will be done such as analyses of subgroups and interactions, and sensitivity analysis if enough data will be available.

## 8. DISCUSSION

In the discussion, we will report the following items:

1. Key results in relation to study objectives.
2. Limitations, considering sources of potential bias or imprecision (we will discuss both the direction and magnitude of any potential bias).
3. A cautious overall interpretation of results considering objectives, limitations, multiplicity of analyses, results from similar studies, and other relevant evidence.
4. Implications of the study.
5. Generalisability (external validity).

## 9. PROJECT TIMELINE

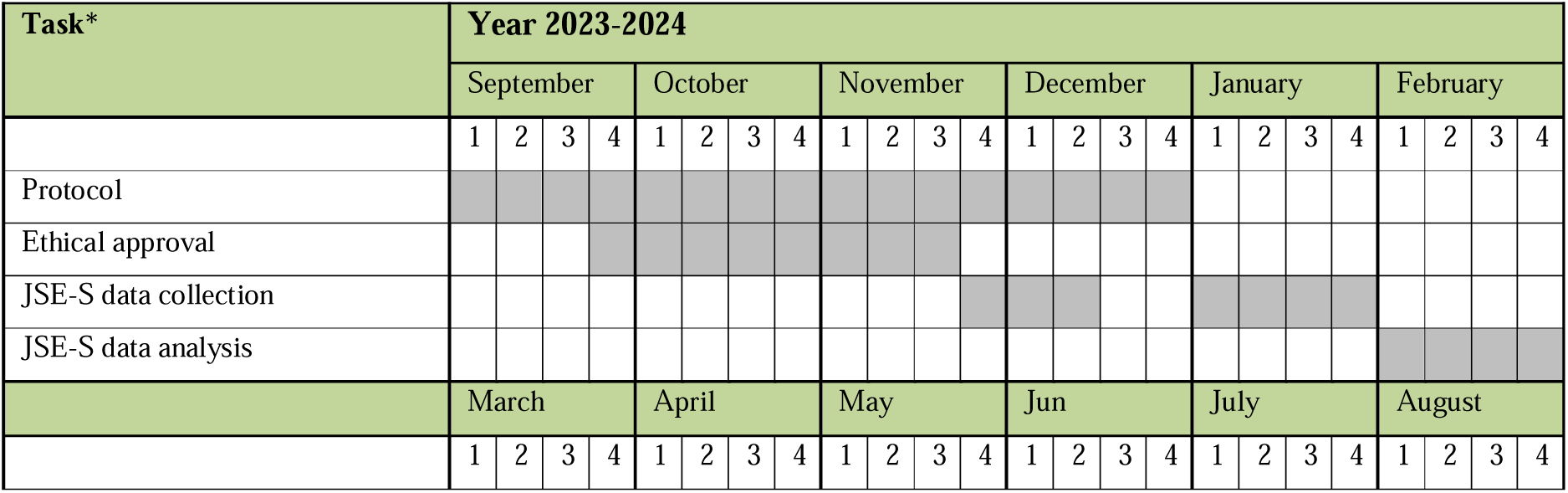
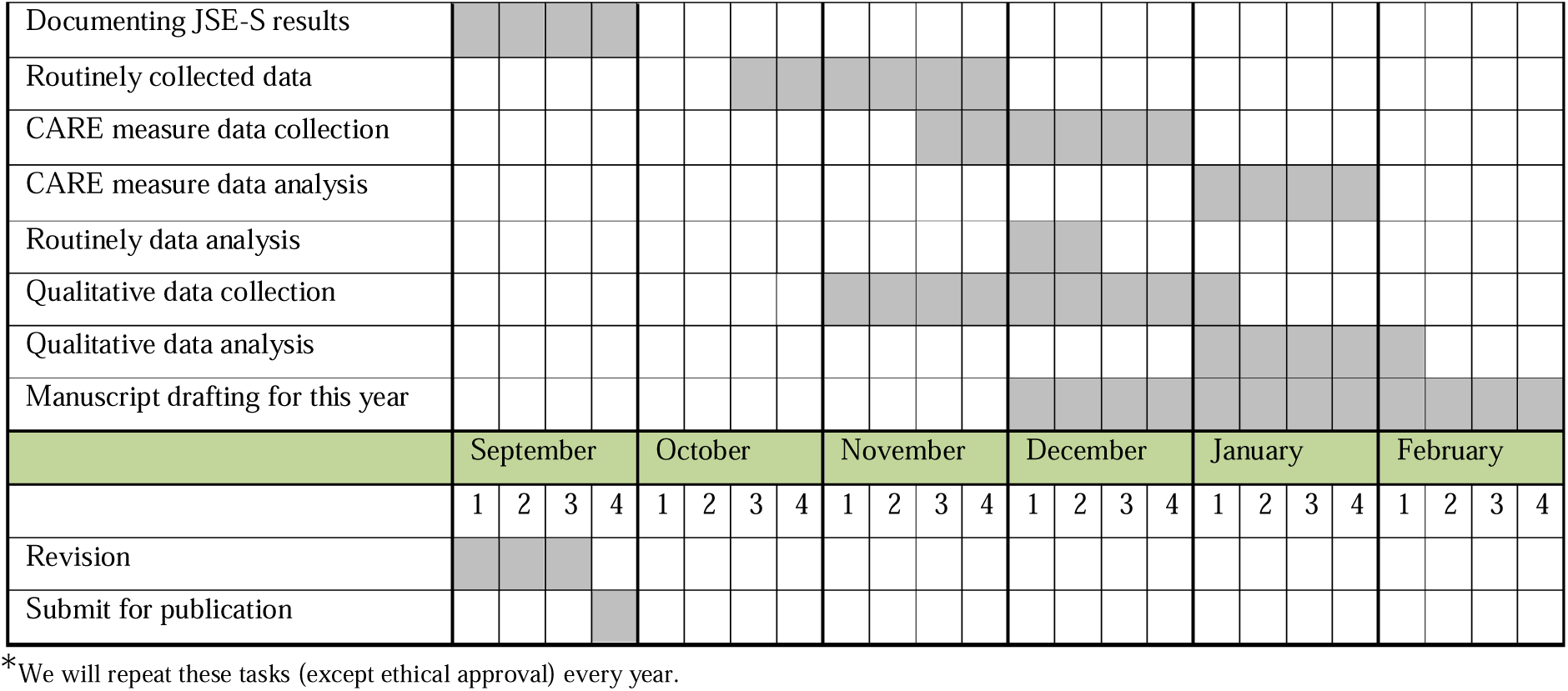

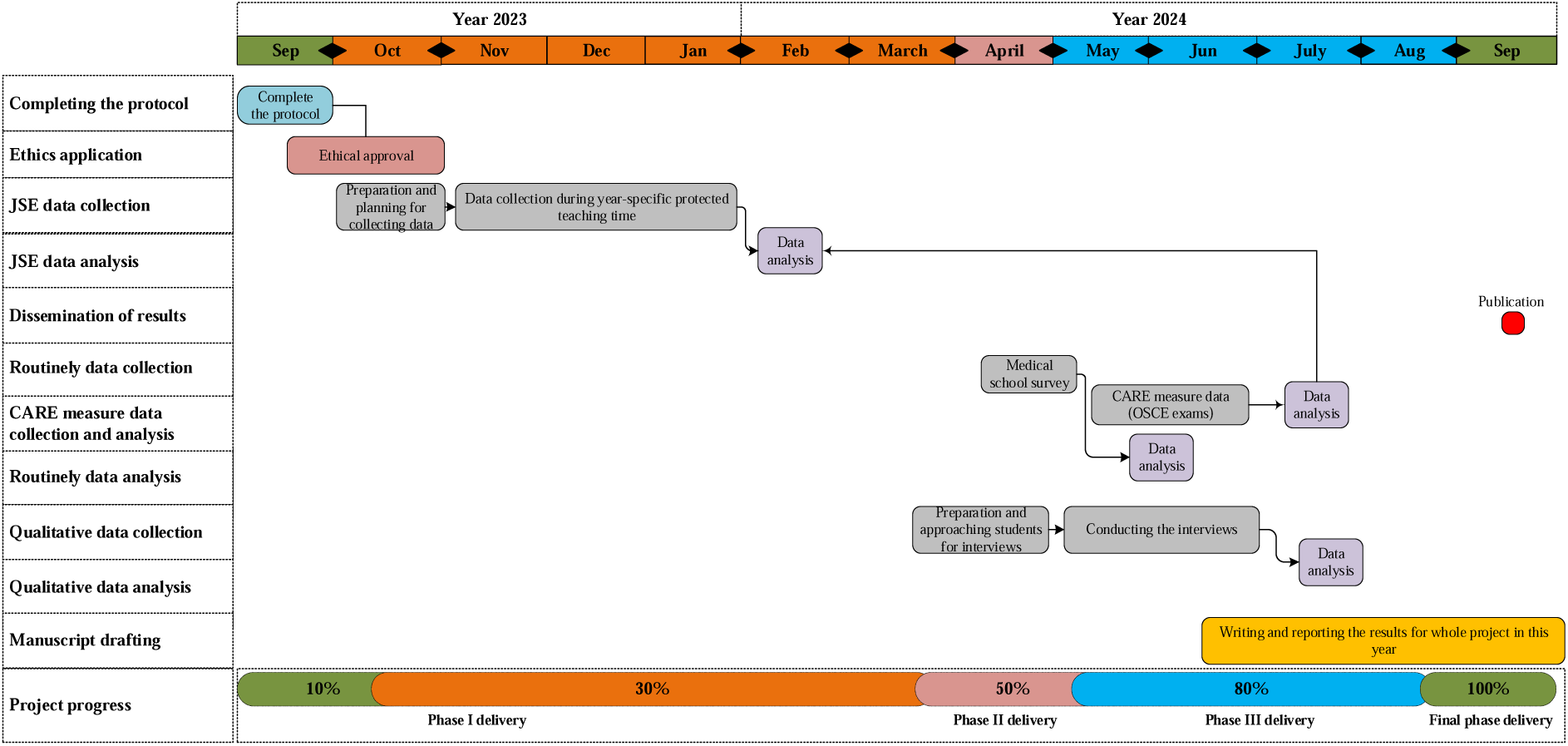

## Data Availability

All data produced in the present study are available upon reasonable request to the authors

## LIST OF ABBREVIATIONS

CARE: Consultation and Relational Empathy
CHDD: Compassionate Holistic Diagnostic Detectives
JSE-S: Jefferson Scale for Empathy Students

