## Supplementary files for "Impact of a novel comprehensive empathy curriculum at Leicester Medical School: Protocol for a longitudinal study"

### Supplement File 1. Interview questions

**Part A. Demographics**

- For the record purpose, could you please state your ethnic group, gender and the year of study?

**Part B. Understanding the Empathy Curriculum**

- Could you please explain what you know about the empathy curriculum at Leicester Medical School? (Based on the year of study)

**Probe**: Can you tell me what teaching you received this year as part of the empathy curriculum?

[Over this past academic year, the empathy curriculum consisted of a description of the empathy curriculum for that academic year, gathered from Andy Ward, and Jeremy Howick.]

**Part C. Teaching**

- Thinking about the empathy curriculum that we discussed above, how engaging did you find the empathy curriculum?

**Probe**: Could you provide specific examples of teaching that you found particularly engaging or disengaging?

- Can you tell me about any memorable or enjoyable teaching sessions within the empathy curriculum?

**Probe**: What made these memorable or enjoyable?

**Part D. Real-world application**

- Could you provide examples where you've been able to apply the empathy skills you've learned in real-world medical scenarios? (Students in year 3, 4 and 5)
- What are the challenges to applying empathy skills in real-world scenarios? (Students in year 3, 4 and 5)

**Probe**: Could you provide examples/more details?

- Have you noticed any changes in your perspective, attitudes, or behaviours since completing the empathy curriculum?

**Probe**: Could you provide examples/explain more?

**Part E. Feedback and improvement**

- To what extent do you think the empathy curriculum aligns with other areas of your medical training?

**Probe**: Could you provide examples/explain more?

**Probe**: How could this be improved?

- Are there any aspects of the curriculum that you feel are missing or could be improved?

**Probe**: Could you provide examples/explain more?

- Based on your experiences so far, what recommendations do you have for refining or enhancing the empathy curriculum for future students?

**Probe**: Is there anything that worked particularly well that we should carry forward?

**Probe:** Is there anything you think we should change or add to improve the curriculum for future students?

**Probe**: Could you provide examples/explain more?

### Supplement File 2. JSE Information Sheet

Research Project title

Impact of a novel comprehensive empathy curriculum at Leicester Medical School: a longitudinal study.

Invitation paragraph

You are being invited to take part in a research project. Before you decide whether or not to take part, it is important for you to understand why the research is being done and what it will involve. Please take time to read the following information carefully.

The data collected as part of this study may be used, in part or in whole, for the writing of research articles, at no time would any personally identifiable data be published without consent.

What is the purpose of the research project?

The Stoneygate Centre for Empathic Healthcare is working with Leicester Medical School to provide students with the opportunity to hone their empathic skills and abilities to an advanced level. This study aims to evaluate the impact of the enhanced empathy curriculum throughout all five years of medical school. To achieve this aim, we will conduct a longitudinal study within Leicester Medical School using both quantitative data (e.g., empathy levels, student satisfaction with the empathy curriculum, satisfaction with the overall medical school curriculum) and qualitative data (about student experiences with the medical school curriculum). We will collect this data every year for 10 years.

Why have I been invited to participate?

All medical students in Leicester Medical School from year one to five are eligible to take part in this study. If you are a first- or second-year student you will be approached about taking part in this survey during your CHDD module teaching. If you are a third, fourth- or fifth-year student you will hear more about this survey during your study days at Leicester Medical School.

Do I have to take part?

It is up to you to decide whether or not to take part in this research project. If you do decide to take part you will be given this information sheet along with a privacy notice that will explain how your data will be collected and used, and be asked to provide your consent to participate. If you decide to take part you are still free to withdraw at any time and without giving a reason, by contacting the researcher.

Please note that choosing not to take part or not will not affect your involvement with the medical school in any way. Participation in this project also will not affect the outcome in your medical degree in any way.

What will happen to me if I take part?

You are being asked to complete a self-rating questionnaire to assess empathy during your medical degree at Leicester Medical School. The results will be anonymised and used for research and evaluation purposes. The questionnaire asks you to rate how strongly you agree or disagree with 20 statements that are designed to measure empathy. It should take no more than 15 minutes to complete.

What are the possible disadvantages and risks of taking part? (where appropriate)

There are no anticipated risks to participating in this research.

What are the possible benefits of taking part?

There are no direct benefits to you for your participation in this study. However, we hope that the information obtained from this study will improve the empathy curriculum for future cohorts.

What data will you collect about me?

The questionnaire asks you to rate how strongly you agree or disagree with 20 statements that are designed to measure empathy. We will not collect any identifying information from you.

Will what I say in this research project be kept confidential?

Your responses to this survey will be anonymous and coded our research team for the purpose of data analysis and confidentiality.

Participant data will be kept confidential except in cases where the researcher is legally obligated to report specific incidents. These incidents include, but may not be limited to, incidents of abuse and suicide risk.

How will you look after the data you collect about me?

We need to ensure that you understand what will happen to the data we collect about you as well as your legal rights. This document is accompanied with a separate Privacy Notice providing further details, you can access this via [insert method here – link to website or paper copy]. You can stop being part of the research project at any time, without giving a reason, but we will keep information about you that we already have and continue to use this for the purposes of the research project as outlined here.

Data will be stored on a secure server in a folder to which only the study researchers have access. Information that you provide and your answers to the survey will be anonymised. At all times this research study will comply with the UK General Data Protection Regulations (2018).

What will happen to the results of the research project?

The results will be disseminated through various publications. A copy of the findings of the research project will be provided to each participant if they would like it.

What should I do if I want to take part?

You will be asked to complete an Informed Consent Form by ticking the Yes. This will confirm you understand how your data will be processed, protected and reviewed for research purposes.

Who is organising and funding the research project?

This study was funded by the Stoneygate Trust. The funder had no role in any part of the development of this project.

What if something goes wrong?

There are no anticipated risks to participating in this research. In the very unlikely event of you being harmed by taking part in this research project, there are no special compensation arrangements. If you are harmed due to someone’s negligence, then you may have grounds for legal action but you may have to pay for it.

Who has reviewed the research project?

This project and associated documents have been approved by the University of Leicester Research Ethics Committee. Please don't hesitate to contact the study lead (Dr Leila Keshtkar on) If you have any further questions about this study.

If you have any concerns or queries about the way in which this project has been conducted, then you should contact the Chair of the University Research Ethics Committee on. If you require more GDPR data protection information then you can access this via the University’s Information Assurance Services:

Information Assurance Services

University of Leicester

University Road

Leicester

LE1 7RH

T: +44 (0)116 229 7945

E:

W: <https://www2.le.ac.uk/offices/ias>

Thank you.

### Supplement File 3. JSE Consent Form

**Full title of Project: Impact of a novel comprehensive empathy curriculum at Leicester Medical School: a longitudinal study**

**Name, position and contact details of Researcher:**

**Leila Keshtkar,** **Research fellow at Stoneygate Centre for Empathic Healthcare,**

**Rachel Winter,** **Associate Professor for Empathic Healthcare and Medical Education,**

**Andy Ward,** **Associate Professor of Medical Education,** ****

**Amber Bennett-Weston,** **Research fellow at Stoneygate Centre for Empathic Healthcare,** ****

**Name, position and contact details for Supervisor: Professor Jeremy Howick,** **Professor of Empathic Healthcare and Director of the Stoneygate Centre for Empathic Healthcare,** ****

|  | Please **initial** box |
| --- | --- |
| 1. I confirm that I have read and understand the participant information sheet (**Version 1, 08 November 2023**) for the above study and have had the opportunity to ask questions. |  |
| 2. I understand that my participation is voluntary and that I am free to withdraw at any time, without giving reason. |  |
| 3. I understand that at all times this research project will comply with the *General Data Protection Regulations (GDPR, 2018)* approved by the EU parliament on 14 April 2016 and passing into UK law effective from 25 May 2018 and that if I have any concerns how I contact the University of Leicester to raise these.   4. I agree to take part in the above research project. |  |

Name of Participant	Date	Signature

Name of Researcher obtaining informed consent	Date	Signature

### Supplement File 4. Privacy Notice for Participants

Research Study title & Researcher Names

Impact of a novel comprehensive empathy curriculum at Leicester Medical School: a longitudinal study.

Leila Keshtkar, Research fellow at Stoneygate Centre for Empathic Healthcare,

Amber Bennett-Weston, Research fellow at Stoneygate Centre for Empathic Healthcare,

Rachel Winter, Associate Professor for Empathic Healthcare and Medical Education,

Andy Ward, Associate Professor of Medical Education,

Jeremy Howick, Professor of Empathic Healthcare and Director of the Stoneygate Centre for Empathic Healthcare,

This Privacy Notice provides information about how the University of Leicester collects and uses your personal information when you take part in this research projects.

Please also refer to the Participant Information Sheet given to you for further details about the research project, what information will be collected about you, and how it will be used.

The University of Leicester will usually be the *Data Controller* of any data that you supply for this research. This means that we are responsible for looking after your information and using it properly. This means that the University will make the decisions on how your data is used and for what reasons. The exception to this is joint research projects, if this is applicable you will be informed on the Participant Information Sheet as to the other partner institution(s) who will also have responsibilities for looking after your information. You can access more information on this via the University’s Information Assurance Services:

Information Assurance Services
University of Leicester
University Road
Leicester
LE1 7RH
T: +44 (0)116 229 7945
E:

W: <https://www2.le.ac.uk/offices/ias>

Why do we need your data?

This study aims to evaluate the impact of the enhanced empathy curriculum throughout all five years of medical school.

University of Leicester’s legal basis for collecting this data is:

Processing is necessary for the performance of a task in the public interest such as research.

What type of data will the University of Leicester use?

This study uses both quantitative data (about empathy levels, student satisfaction with the empathy curriculum, satisfaction with the overall medical school curriculum and medical school average grades) and qualitative data (about student experiences with the medical school curriculum).

Who will the University of Leicester share your data with?

Not applicable.

Will the University of Leicester transfer my data outside of the UK?

Not applicable.

What rights do I have regarding my data held by the University of Leicester?

Your normal rights under the Data Protection Act and the General Data Protection Regulation apply. However, we need to manage your records in specific ways for the research project to be reliable. This means that we will not [always] be able to let you see or change the data we hold about you.

You can stop being part of the research project at any time, without giving a reason, but we will keep information about you that we already have and continue to use this for the purposes of the research project as outlined in the Participant Information Sheet.

Where did the University of Leicester source my data from?

Participants

Are there any consequences of not providing the requested data?

There are no consequences of not providing data for this research. It is purely voluntary.

Will there be any automated decision making using my data?

There will be no use of automated decision making in scope of UK Data Protection and Privacy legislation.

How long will the University of Leicester keep my data?

In line with the law, we will only keep your data for as long as we need to so that we can fulfil our research objectives.

We will keep anonymised research data we have generated for [10 years], this is due to [this a longitudinal study].

Who can I contact if I have concerns?

In the event of any questions about the research project, please contact the researchers in the first instance. The email address is: Dr Leila Keshtkar on

If you have any concerns about the way in which the research project has been conducted, please contact the Chair of the University Research Ethics Committee at.

The University of Leicester Data Protection Officer is:

Data Protection Officer

University of Leicester,

University Road, Leicester, LE1 7RH

0116 229 7640

For further details about information security, please contact the [Information Assurance Services](https://www2.le.ac.uk/offices/ias) team.

### Supplement File 5. CARE Information Sheet

### Supplement File 6. CARE Consent Form

### Supplement File 7. Interview Information Sheet

Research Project title

Impact of a novel comprehensive empathy curriculum at Leicester Medical School: a longitudinal study.

Invitation paragraph

You are being invited to take part in a research project. Before you decide whether or not to take part, it is important for you to understand why the research is being done and what it will involve. Please take time to read the following information carefully.

The data collected as part of this study may be used, in part or in whole, for the writing of research articles, at no time would any personally identifiable data be published without consent.

What is the purpose of the research project?

The Stoneygate Centre for Empathic Healthcare is working with Leicester Medical School to provide students with the opportunity to hone their empathic skills and abilities to an advanced level. This study aims to evaluate the impact of the enhanced empathy curriculum throughout all five years of medical school. To achieve this aim, we will conduct a longitudinal study within Leicester Medical School using both quantitative data (e.g., empathy levels, student satisfaction with the empathy curriculum, satisfaction with the overall medical school curriculum) and qualitative data (about student experiences with the medical school curriculum). We will collect this data every year for 10 years.

Why have I been invited to participate?

All medical students in Leicester Medical School from year one to five are eligible to take part in this study. If you are a first- or second-year student you will be approached about taking part in this survey during your CHDD module teaching. If you are a third, fourth- or fifth-year student you will hear more about this survey during your study days at Leicester Medical School.

Do I have to take part?

It is up to you to decide whether or not to take part in this research project. If you do decide to take part you will be given this information sheet along with a privacy notice that will explain how your data will be collected and used, and be asked to provide your consent to participate. If you decide to take part you are still free to withdraw at any time and without giving a reason, by contacting the researcher.

Please note that choosing not to take part or not will not affect your involvement with the medical school in any way. Participation in this project also will not affect the outcome in your medical degree in any way.

What will happen to me if I take part?

You are being asked to complete a self-rating questionnaire to assess empathy during your medical degree at Leicester Medical School. The results will be anonymised and used for research and evaluation purposes. The questionnaire asks you to rate how strongly you agree or disagree with 20 statements that are designed to measure empathy. It should take no more than 15 minutes to complete.

What are the possible disadvantages and risks of taking part? (where appropriate)

There are no anticipated risks to participating in this research.

What are the possible benefits of taking part?

There are no direct benefits to you for your participation in this study. However, we hope that the information obtained from this study will improve the empathy curriculum for future cohorts.

What data will you collect about me?

The questionnaire asks you to rate how strongly you agree or disagree with 20 statements that are designed to measure empathy. We will not collect any identifying information from you.

Will what I say in this research project be kept confidential?

Your responses to this survey will be anonymous and coded our research team for the purpose of data analysis and confidentiality.

Participant data will be kept confidential except in cases where the researcher is legally obligated to report specific incidents. These incidents include, but may not be limited to, incidents of abuse and suicide risk.

How will you look after the data you collect about me?

We need to ensure that you understand what will happen to the data we collect about you as well as your legal rights. This document is accompanied with a separate Privacy Notice providing further details, you can access this via [insert method here – link to website or paper copy]. You can stop being part of the research project at any time, without giving a reason, but we will keep information about you that we already have and continue to use this for the purposes of the research project as outlined here.

Data will be stored on a secure server in a folder to which only the study researchers have access. Information that you provide and your answers to the survey will be anonymised. At all times this research study will comply with the UK General Data Protection Regulations (2018).

What will happen to the results of the research project?

The results will be disseminated through various publications. A copy of the findings of the research project will be provided to each participant if they would like it.

What should I do if I want to take part?

You will be asked to complete an Informed Consent Form by ticking the Yes. This will confirm you understand how your data will be processed, protected and reviewed for research purposes.

Who is organising and funding the research project?

This study was funded by the Stoneygate Trust. The funder had no role in any part of the development of this project.

What if something goes wrong?

There are no anticipated risks to participating in this research. In the very unlikely event of you being harmed by taking part in this research project, there are no special compensation arrangements. If you are harmed due to someone’s negligence, then you may have grounds for legal action but you may have to pay for it.

Who has reviewed the research project?

This project and associated documents have been approved by the University of Leicester Research Ethics Committee. Please don't hesitate to contact the study lead (Dr Leila Keshtkar on) If you have any further questions about this study.

If you have any concerns or queries about the way in which this project has been conducted, then you should contact the Chair of the University Research Ethics Committee on. If you require more GDPR data protection information then you can access this via the University’s Information Assurance Services:

Information Assurance Services

University of Leicester

University Road

Leicester

LE1 7RH

T: +44 (0)116 229 7945

E:

W: <https://www2.le.ac.uk/offices/ias>

Thank you.

### Supplement File 8. Interview Consent Form

**Full title of Project: Impact of a novel comprehensive empathy curriculum at Leicester Medical School: a longitudinal study**

**Name, position and contact details of Researcher: Leila Keshtkar,** **Research fellow at Stoneygate Centre for Empathic Healthcare,**

**Rachel Winter,** **Associate Professor for Empathic Healthcare and Medical Education,**

**Andy Ward,** **Associate Professor of Medical Education,** ****

**Amber Bennett-Weston,** **Research fellow at Stoneygate Centre for Empathic Healthcare,** ****

**Name, position and contact details for Supervisor: Professor Jeremy Howick,** **Professor of Empathic Healthcare and Director of the Stoneygate Centre for Empathic Healthcare,** ****

|  | Please **initial** box |
| --- | --- |
| 2. I confirm that I have read and understand the participant information sheet (**Version 1, 09 November 2023**) for the above study and have had the opportunity to ask questions. |  |
| 4. I understand that my participation is voluntary and that I am free to withdraw at any time, without giving reason. |  |
| 5. I understand that at all times this research project will comply with the *General Data Protection Regulations (GDPR, 2018)* approved by the EU parliament on 14 April 2016 and passing into UK law effective from 25 May 2018 and that if I have any concerns how I contact the University of Leicester to raise these.   4. I agree to take part in the above research project. |  |
|  | Yes No |
| 5. I understand that the interview will be audio-recorded. |  |
| 6. I agree to the use of anonymised quotes in publications. |  |

Name of Participant	Date	Signature

Name of Researcher obtaining informed consent	Date	Signature
